# Sex differences in depression and sleep disturbance as inter-related risk factors of diabetes

**DOI:** 10.1101/2022.05.20.22275367

**Authors:** Clara S. Li, Rose Porta, Shefali Chaudhary

## Abstract

**Background:** Previous studies identified depression and sleep disturbance as risk factors for diabetes. Sleep disturbance and depression are known to be inter-related. Further, women relative to men are more prone to depression. Here, we investigated how depression and sleep disturbance may jointly influence the risk of diabetes and the effects of sex on these influences.

**Methods:** Using the data of 21,229 participants from the 2018 National Health Interview Survey, we performed multivariate logistic regression with diabetes diagnosis as the dependent variable, sex, self-reported frequency of weekly depression and nightly sleep duration, and their interactions with sex as independent variables, and age, race, income, body mass index and physical activity as covariates. We employed Bayesian and Akaike Information criteria to identify the best model, evaluated the accuracy of the model in predicting diabetes using receiver operating characteristic analysis, and computed the odds ratios of these risk factors.

**Results:** In the two best models, depression frequency and sleep hours interact with sex in determining the diagnosis of diabetes, with higher depression frequency and nightly duration of sleep longer or shorter than 7 to 8 hours associated with higher likelihood of diabetes. The two models both predicted diabetes at an accuracy (area under the receiver operating characteristic curve) of 0.86. Further, these effects were stronger in men than in women at each depression and sleep level.

**Conclusions:** Depression and sleep inter-relatedly rather than independently contributes to diabetes. Depression and sleep hours associate with diabetes more significantly in men than in women. The current findings indicate a sex-dependent relationship between depression, sleep disturbance and diabetes risk and add to a growing body of evidence linking mental and physical health.

## INTRODUCTION

Over 34 million Americans, or more than 1 in 10, are diagnosed with diabetes, one of the most prevalent chronic diseases in the US, according to the Centers for Disease Control and Prevention, National Diabetes Statistics Report, 2020. Diabetes and related health problems have an enormous impact on US health care spending. Individuals with diabetes face an overall 60% greater risk of early death than non-diabetics, underscoring the importance of identifying risk factors for targeted treatment [1].

In addition to diet and physical activity, sleep and mental health have been investigated as modifiable risk factors of diabetes [2]. Studies examining the relationship between sleep and diabetes associate both short (<6 h) and long (>8 h) sleep duration with increased diabetes risk as strongly as overweight and family history of diabetes [3]. Depression has also been associated with higher risk for diabetes. For instance, a meta-analysis reported that individuals with depression are 37% more likely to develop type 2 diabetes than their depression-free counterparts [4]. However, sleep disturbance is one of the core manifestations of depression [5–7]. Individuals with depression can demonstrate insomnia or hypersomnia along with other physical symptoms. Thus, whereas sleep and depression have largely been characterized as independent risk factors for diabetes, it is highly likely that depression and sleep disturbance conduce to diabetes in an inter-related manner.

It is known that women relative to men are more vulnerable to depression, as demonstrated in a meta-analysis [8]. The latter study showed that sex differences in the risk of depression emerges early in the life span with an odds ratio (OR) peaking at adolescence (OR=3.02) and declining afterwards and remaining stable in adulthood (OR=1.71 to 2.02). Thus, about twice as many female experience depression as male adults. Many studies have investigated the genetic [8], neurobiological [9,10] and socio-psychological [11] mechanisms underlying sex differences in the prevalence of depression. Indeed, women have more sleep-related complaints, including insomnia, than men [12] and particularly so in older age [13]. Thus, it is highly likely that diabetes may relate to depression and sleep disturbance differently between the sexes [14].

Here, using a large public dataset we investigated how depression and sleep disturbance relate to diabetes and whether the relationships vary between women and men. Based on the literature, we hypothesized that greater severity of depression would be associated with higher likelihood of diabetes and that nightly sleep duration would be associated with diabetes in a U-shaped function, with short and long hours of sleep associated with higher risk. Further, these relationships would be stronger in females than in males.

## METHODS

### Data set and variables

We used the Integrated Public Use Microdata Series (IPUMS) National Health Interview Survey Data, 2018, of 21,229 participants. The data were collected by random sampling of 35,000 U.S. households and random selection of one adult and one child (if any) from each household for interview. We restricted the sample to those 18 years or older as children and adolescents demonstrate sleep patterns and durations different from those of adults. We intended to generalize the current findings to non-institutionalized adult population of the United States.

The variables in our analytics included a binary response (dependent) variable indicating whether an individual has diabetes (the IPUMS data set did not distinguish between type I or II of diabetes). Independent variables included 1) the average number of self-reported hours of sleep per night over the prior month in four categories: ≤ 5, 6-7, 7-8, and >8 hours; 2) the frequency an individual experiencing depression over the prior year in three categories: never/rarely, monthly, and often (more often than monthly); and 3) sex: males and females. Covariates included age (in years), race (Caucasian, black/African American, Asian, or Native American/Alaska Native), income, body mass index (BMI), and level of physical activity (PA). Household income was categorized into low (annual family income < $35,000), middle (between $35,000 and $75,000), and high (> $75,000). The variable measuring PA was created using the Oncology Nursing Society formula for weekly leisure activity score, derived from the Godin Leisure-Time Exercise Questionnaire. The data for ‘light exercise’ were not available and thus substituted with the data on strength-related exercise. **Table 1** shows the demographic and clinical characteristics of the sample.

**Table 1.** Demographic and clinical characteristics of the sample

|  | **Men (n=9,860, 46.4%)** |  | **Women (n=11,369, 53.6%)** |  |
| --- | --- | --- | --- | --- |
|  | **Diabetes (+)** | **Diabetes (-)** | **Diabetes (+)** | **Diabetes (-)** |
| **N (%)** | 198 (2.00%) | 9,662 (98.00%) | 209 (1.84%) | 11,160 (98.16%) |
| **Age (yr, mean ± SD)** | 65.3 ± 11.6 | 49.8 ± 17.9 | 65.4 ± 12.6 | 51.2 ± 18.4 |
| **BMI (mean ± SD)** | 32.4 ± 8.3 | 28.3 ± 5.6 | 33.1 ± 8.3 | 27.9 ± 6.8 |
| **PA, median (range)** | 1615 (45-1615) | 796 (29-1615) | 1615 (68-1615) | 1145 (31-1615) |
| **Race** |  |  |  |  |
| Caucasian | 153 (1.89%) | 7,939 (98.11%) | 131 (1.43%) | 8,992 (98.56%) |
| Asian | 10 (1.76%) | 558 (98.24%) | 13 (2.16%) | 588 (97.84%) |
| African American | 25 (2.36%) | 1,035 (97.64%) | 59 (3.98%) | 1,423 (96.02%) |
| Native American | 10 (7.14%) | 130 (92.86%) | 6 (3.68%) | 157 (96.32%) |
| **Income** |  |  |  |  |
| Low | 105 (3.68%) | 2,744 (96.31%) | 149 (3.61%) | 3,977 (96.39%) |
| Middle | 63 (2.16%) | 2,854 (97.84%) | 36 (1.14%) | 3,120 (98.86%) |
| High | 30 (0.73%) | 4,064 (99.27%) | 24 (0.59%) | 4,063 (99.41%) |
| **Depression** |  |  |  |  |
| Rarely/never | 133 (1.57%) | 8,342 (98.43%) | 138 (1.50%) | 9,073 (98.50%) |
| Monthly | 17 (3.33%) | 493 (96.67%) | 17 (1.96%) | 849 (98.04%) |
| Often | 48 (5.48%) | 827 (94.5%) | 54 (4.18%) | 1,238 (95.82%) |
| **Sleep hours/night** |  |  |  |  |
| $\leq 5$ | 31 (3.33%) | 900 (96.67%) | 40 (3.41%) | 1,132 (96.59%) |
| 6-7 | 50 (2.19%) | 2,237 (97.81%) | 49 (1.88%) | 2,554 (98.12%) |
| 7-8 | 76 (1.30%) | 5,784 (98.70%) | 84 (1.27%) | 6,516 (98.73%) |
| >8 | 41 (5.24%) | 741 (94.76%) | 36 (3.62%) | 958 (96.38%) |

Note: BMI: body mass index; PA: physical activity

### Statistical analysis

#### Multivariate logistic regression

We used binomial logistic regression to predict binary outcome (presence/absence) of diabetes using an optimized model [15]. The variables of interest included sex, depression frequency (depression), and nightly hours of sleep (sleep), and the covariates included age, race, household income (income), BMI, and PA.

We first performed a multivariate logistic regression to include all predictors (sex, frequency of depression, and nightly duration of sleep) and covariates (age, race, BMI, PA, and income) in the model. We checked for the statistical significance of the model; further, the three predictors and five covariates showed a significant relationship with diabetes and thus were all included in the model (see Results).

Next, we examined the continuous variables (age, BMI, and PA) for linearity in relation to the logit of the outcome. The results showed that age, BMI and PA were all linearly related to diabetes in logit scale (**Figure 1**).

**Figure 1.**
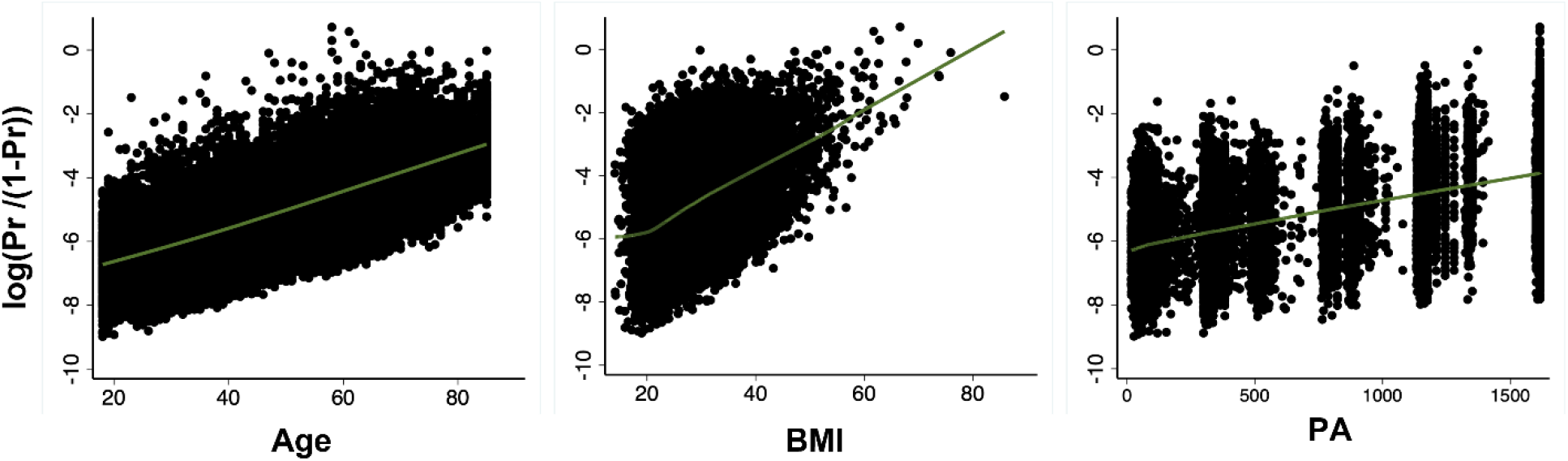
Scatter plots with locally weighted scatter plot smoothing (LOWESS) of the relationship between probability of diabetes (in logit scale) and continuous covariates (age, BMI, PA, respectively). All relationships were linear.

#### Identification of prediction model with the best fit

The findings of multivariate logistic regression showed that all 3 predictors and 5 covariates showed significant relationships with diabetes (**Table 2**). The goal of the study was to investigate how depression frequency, sleep duration and sex may interact in the contribution to diabetes. Thus, we employed Bayesian Information Criterion (BIC) and Akaike Information Criterion (AIC) to determine which of the following models showed the best fit:

**Table 2.** Multivariate logistic regression

| Variable | Coefficient (SE) | Odds ratio (SE) | p-value |
| --- | --- | --- | --- |
| Sleep (hours/night; 7-8 hours/night as ref.) |  |  |  |
| ≤5 | 0.49 (0.15) | 1.6 (0.25) | 0.001 |
| 6-7 | 0.35 (0.13) | 1.4 (0.19) | 0.008 |
| >8 | 0.49 (0.15) | 1.6 (0.24) | 0.001 |
| **Depression frequency (Never/Rarely as ref.)** |  |  |  |
| Monthly | 0.59 (0.19) | 1.8 (0.35) | 0.002 |
| Often | 0.84 (0.13) | 2.3 (0.30) | <0.001 |
| **Sex (Men as ref.)** | -0.47 (0.11) | 0.6 (0.07) | <0.001 |
| **Age** | 0.05 (0.004) | 1.1 (0.004) | <0.001 |
| **BMI** | 0.08 (0.006) | 1.1 (0.007) | <0.001 |
| **PA** | 0.0007 (0.001) | 1.0 (0.0001) | <0.001 |
| **Race (Caucasian as ref.)** |  |  |  |
| Asian | 0.88 (0.23) | 2.4 (0.55) | <0.001 |
| African American | 0.59 (0.13) | 1.8 (0.24) | <0.001 |
| Native American | 1.18 (0.28) | 3.2 (0.91) | <0.001 |
| **Income (Low as ref.)** |  |  |  |
| Medium | -0.53 (0.13) | 0.59 (0.07) | <0.001 |
| High | -0.95 (0.16) | 0.37 (0.06) | <0.001 |

**Note:** Ref., reference; SE, standard error

*Model 1*: Diabetes = depression + sleep + sex + sex × depression + age + BMI + PA + race + income

*Model 2*: Diabetes = depression + sleep + sex + sex × sleep + age + BMI + PA + race + income

*Model 3*: Diabetes = depression + sleep + sex + depression × sleep + age + BMI + PA + race + income

*Model 4*: Diabetes = depression + sleep + sex + sex × depression × sleep + age + BMI + PA + race + income

*Model 5*: Diabetes = depression + sleep + sex + sex × depression + sex × depression × sleep + age + BMI + PA + race + income

*Model 6*: Diabetes = depression + sleep + sex + sex × sleep + sex × depression × sleep + age + BMI + PA + race + income

*Model 7*: Diabetes = depression + sleep + sex + depression × sleep + sex × depression × sleep + age + BMI + PA + race + income

*Model 8*: Diabetes = depression + sleep + sex + sex × depression + sex × sleep + depression × sleep + sex × depression × sleep + age + BMI + PA + race + income

Model fit according to BIC and AIC was assessed using Hosmer and Lemeshow’s goodness-of-fit test [16].

#### Consideration of influential observations

Logistic regression is highly sensitive to the presence of influential observations (outliers). We assessed potential outliers and their impact on the model. We computed model’s standardized Pearson’s residuals, adjusted for the number of observations that shared the same covariate pattern, to detect potential outliers (i.e., data points with large deviations between observed and fitted values) [17]. We employed Pregibon delta beta statistics and Pregibon leverage to evaluate the influence of a data point on estimated coefficients of the model [18]. Pregibon delta beta measures the extent to which inclusion or exclusion of an observation sharing the same covariate pattern changes the beta estimates (coefficient). It is presented in the form of standardized difference in betas, with larger values indicating larger influence on model estimates. Pregibon leverage computes the diagonal elements of the hat matrix adjusted for the number of observations that share the same covariate pattern. Large values indicate covariate patterns farther from the average and a larger effect on the fitted model.

#### Discriminatory power of the model

We evaluated the discriminatory power of the best model using Receiver Operating Characteristic (ROC) analysis [19]. We showed the sensitivity and specificity vs. probability cut-off points and the ROC curve in a plot of sensitivity vs. (1 – specificity).

## RESULTS

### Multivariate logistic regression

**Table 2** shows the results of multivariate logistic regression. For nightly hours of sleep, we used the category of 7-8 hours as the baseline; for depression frequency, we used “never/rarely” as the baseline; for sex, we used men as the baseline; for race, we used Caucasian as baseline; and, finally, for income, we used “low” income as the baseline. Depression reported at a higher frequency and sleep duration either longer or shorter than 7-8 hours both significantly contributed to the odds of a diagnosis of diabetes.

### Model fit according to BIC and AIC

Although the BIC and AIC values were close across models, the two best models were Model 1 and 2, according to both criteria (**Table 3**):

**Table 3.** Bayesian Information Criterion (BIC) and Akaike Information Criterion (AIC) values of *Model 1 – Model 8*

| **Logistic models** | **AIC** | **BIC** |
| --- | --- | --- |
| *Model 1\** | 3308.09 | 3443.46 |
| *Model 2\** | 3311.56 | 3454.89 |
| *Model 3* | 3314.97 | 3482.20 |
| *Model 4* | 3326.73 | 3581.55 |
| *Model 5* | 3326.73 | 3581.55 |
| *Model 6* | 3326.73 | 3581.15 |
| *Model 7* | 3326.73 | 3581.55 |
| *Model 8* | 3326.73 | 3581.55 |

*Model 1:* Diabetes = depression + sleep + sex + sex × depression + age + BMI + PA + race + income, with a BIC value of 3443.46 and AIC value of 3308.09

*Model 2:* Diabetes = depression + sleep + sex + sex × sleep + age + BMI + PA + race + income, with a BIC value of 3454.89 and AIC value of 3311.56

Hosmer and Lemeshow’s goodness of fit revealed non-significant difference between predicted and observed frequencies (Model 1: p=0.792; Model 2: p= 0.342) with Hosmer-Lemeshow χ^2^ = 4.68 (Model 1) and 9.00 (Model 2), respectively, indicating a good fit for both models.

#### Influential observations

We show the scatter plot of standardized Pearson’s residual versus Pregibon leverage in **Supplementary Figure S1**. The scatter plot combined three diagnostic values and revealed 10 data points to be potential outliers according to all three statistics (residuals >2 AND delta beta >0.01 AND leverage >0.01). We thus repeated logistic regression excluding these 10 data points. As shown in **Supplementary Table S1**, the exclusion of these subjects did not materially affect the variable coefficients; thus, we retained and presented findings based on the entire sample.

#### Discriminatory power of Model 1 and 2

In ROC analysis both model 1 and 2 demonstrated excellent discriminatory power distinguishing the presence vs. absence of diabetes, with an area under curve (AUC) = 0.86 (**Figure 2**). In the plot of sensitivity and specificity versus probability cut-offs, the sensitivity and specificity curves crossed at a point close to the vertical axis, indicating adequate sensitivity and specificity of the fitted models [19].

**Figure 2.**
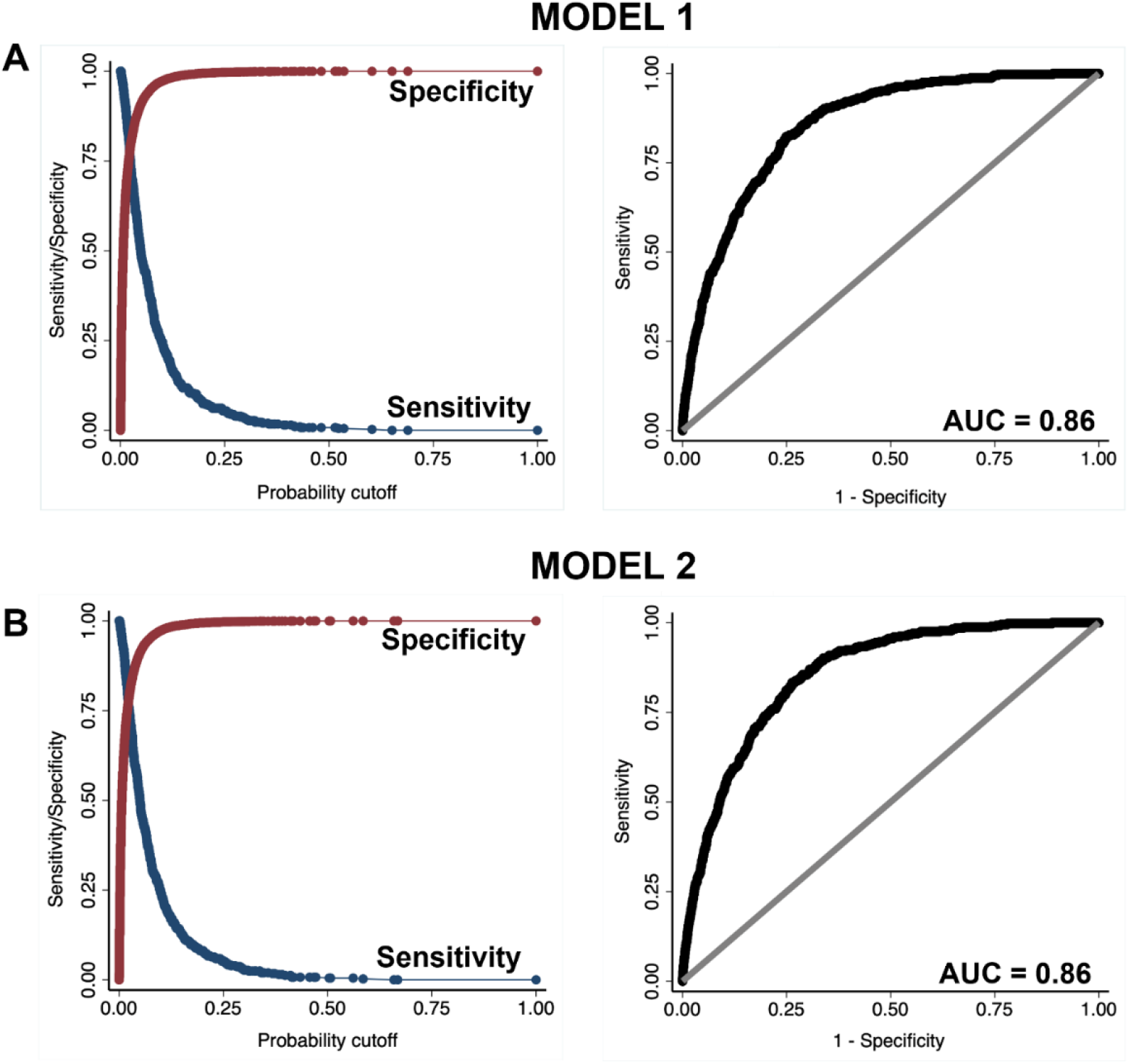
Sensitivity vs. specificity plot (left panels) and receiver operating characteristic (ROC) curve (right panels) of (A) model 1 and (B) model 2.

#### The effects of depression and sleep on the probability of diabetes and their interaction with sex

To visualize the effects of depression frequency and nightly sleep duration on the probability of diabetes, we plotted predictive margins of response (outcome: diabetes) versus predictor in **Figure 3A** and **3B**. Overall, as revealed by logistic regression (**Table 2**), more frequent experience of depression was positively related to diabetes risk. On the other hand, nightly sleep hours were associated with diabetes risk in a U function; duration more or less than 7-8 hours was both associated with a higher risk.

**Figure 3.**
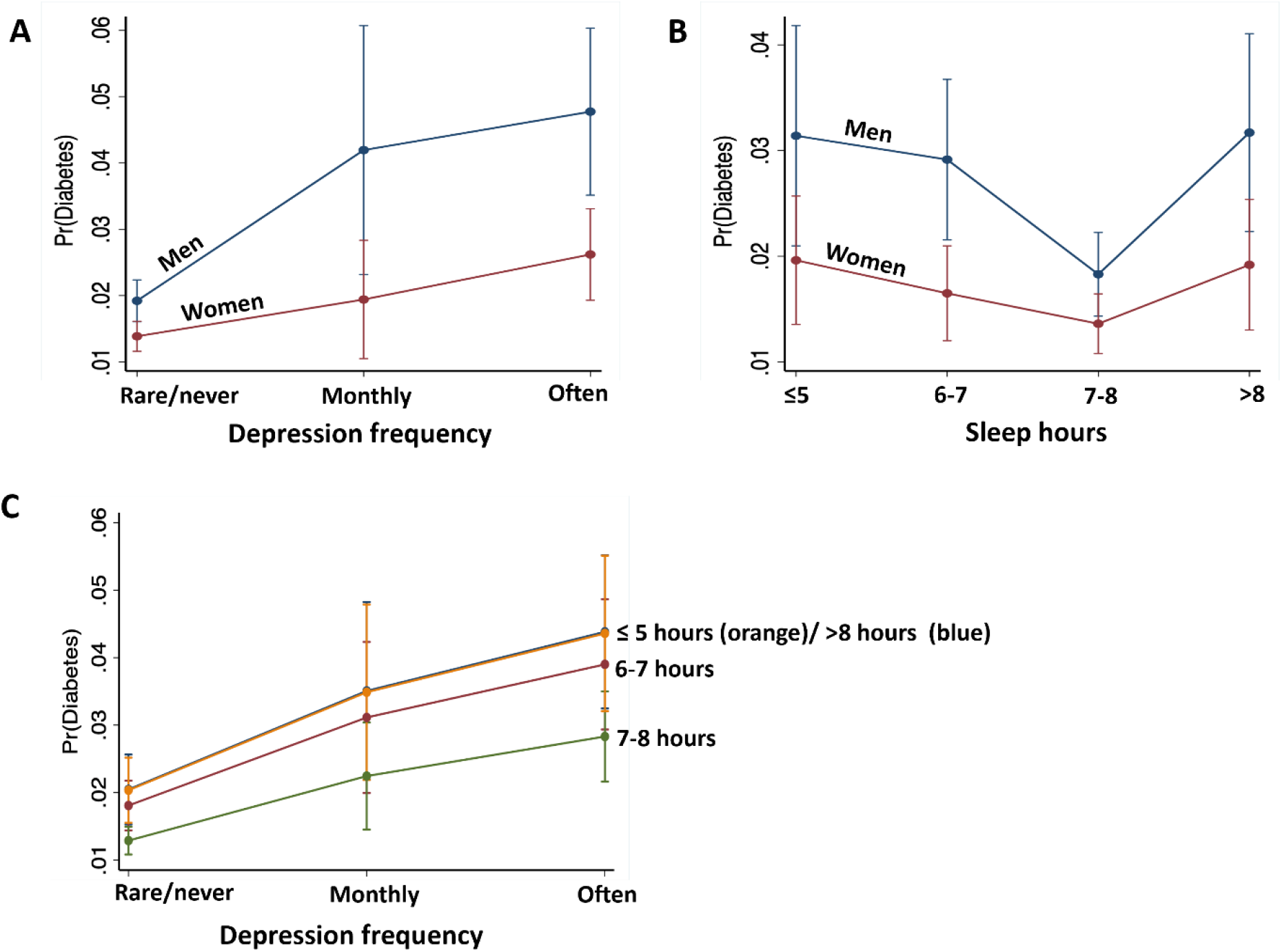
Predicted margins of diabetes with 95% confidence intervals with respect to (A) depression frequency, (B) nightly sleep hours, shown separately for males and females, and (C) depression at each level of sleep. Men relative to women showed stronger associations between depression frequency and nightly sleep hours with the probability of diabetes. Further, reduced/increased sleep hours in synergy with higher frequency of depression predicts higher probability of diabetes.

To examine whether depression frequency and nightly sleep duration may interact to determine diabetes risk, we obtained predictive margins of the interaction effect on diabetes probability using Model 1 (**Figure 3C, Supplementary Table S2**). For each level of sleep hours, diabetes probability increases with more frequent depression in the same manner. Sleep levels ≤5 hours and >8 hours shared similar, highest probability of diabetes at each level of depression, followed by the probability associated with 6-7 and 7-8 hours of sleep, the latter with the least probability of diabetes.

Further, the effects of depression frequency and nightly hours of sleep interacted with sex in predicting diabetes. The effects of depression and sleep disturbance contributed to diabetes more significantly in men than in women (**Table 4**). Specifically, at each level of depression, diabetes probability was higher in men than in women: rarely/never depression [coefficient (SE), odds-ratio (SE), p-value in women with men as reference: -0.35 (0.13), 0.70 (0.09), 0.006], monthly depression [-0.86 (0.37), 0.42 (0.15), 0.019], “often” depression [-0.68 (0.21), 0.50 (0.11), 0.001].

**Table 4.** The effects of depression and sleep interaction with sex on the probability of diabetes (Model 1 and 2)

| Variable | Coefficient (SE) | Odds ratio (SE) | p-value |
| --- | --- | --- | --- |
| **Model 1: Depression × sex (rarely/never depression with men as ref.)** |  |  |  |
| D(rarely/never)s(W) | -0.35 (0.13) | 0.70 (0.09) | 0.006 |
| D(monthly)s(M) | 0.87 (0.28) | 2.39 (0.67) | 0.002 |
| D(monthly)s(W) | 0.01 (0.27) | 1.01 (0.27) | 0.967 |
| D(often)s(M) | 1.02 (0.18) | 2.7 (0.51) | <0.001 |
| D(often)s(W) | 0.34 (0.18) | 1.41 (0.25) | 0.054 |
| **Model 2: Sleep × sex (7-8 hours of sleep of men as ref.)** |  |  |  |
| S(≤5 h)s(M) | 0.59 (0.23) | 1.82 (0.41) | 0.008 |
| S(≤5 h)s(W) | 0.08 (0.21) | 1.07 (0.23) | 0.720 |
| S(5-6 h)s(M) | 0.51 (0.19) | 1.67 (0.32) | 0.007 |
| S(5-6 h)s(W) | -0.11 (0.19) | 0.89 (0.17) | 0.561 |
| S(7-8 h)s(W) | -0.32 (0.16) | 0.73 (0.12) | 0.053 |
| S(>8 h)s(M) | 0.61 (0.21) | 1.84 (0.38) | 0.004 |
| S(>8 h)s(W) | 0.05 (0.22) | 1.05 (0.23) | 0.811 |

D: depression, S: sleep, s: sex, W: women, M: men, SE: standard error.

Similarly, at each level of sleep hours, probability of diabetes was greater in men compared to women: sleep < 5 hours [coefficient (SE), odds-ratio (SE), p-value: -0.52 (0.25), 0.59 (0.15), 0.042], 6-7 hours [-0.62 (0.21), 0.53 (0.11), 0.003)], 7-8 hours [-0.32 (0.16), 0.73 (0.12), 0.053], and >8 hours [-0.55 (0.24), 0.57 (0.14), 0.023].

## DISCUSSION

We assessed how sleep duration and depression frequency related to diabetes diagnosis and how these relationships differed between men and women. Both short (< 7 hours/night) and long (> 8 hours/night) sleep durations were associated with a higher probability of a diagnosis of diabetes. Confirming our first hypothesis, this finding aligns with prior studies documenting a U-shaped relationship between sleep duration and diabetes risk [3,14]. Likewise, experiencing depression daily or weekly was associated with higher probability of a diagnosis of diabetes. Importantly, these relationships were consistent with the symptoms of insomnia and hypersomnia in depression and were stronger in men than in women. Further, the addition of an interaction term of depression × sleep or depression × sleep × sex did not result in better model fit. These findings together suggest an inter-related mechanism associating both depression and sleep disturbance to diabetes. Further, we observed a stronger association in men than in women between these risk factors and diabetes, contrary to our hypothesis. Although the latter findings was largely consistent with the literature, the physiological and psychological mechanisms underlying the sex differences remain to be investigated.

### Sleep disturbance and depression as risk factors of diabetes

We found that, relative to 7 to 8 hours of nightly sleep, both shorter (< 5 hours) and longer (> 8 hours) sleep were associated with an odds ratio of 1.6 of having a diagnosis of diabetes. A systematic review of 36 studies showed that sleep disturbances significantly impact diabetes risk, with the effects comparable to family history of diabetes and overweight and exceeding the risk associated with physical inactivity [1]. A meta-analysis of 13 independent cohorts with a total of 107,756 male and female participants and 3,586 cases of type 2 diabetes reported that both short and long sleep duration was associated with the likelihood of diabetes, although the underlying mechanisms may be different [3]. There is a relatively strong consensus in the literature that short sleep duration (< 6 hours) is a significant risk factor of diabetes [1,20–22]. Insufficient sleep and poor sleep quality may contribute to diabetes and/or hamper treatment via physiologic mechanisms such as insulin resistance, decreased leptin/increased ghrelin, and tissue inflammation as well as behavioral mechanisms, including elevated food intake, smoking, drinking and sedentary behavior, all disposing individuals to both obesity and diabetes [23]. Indeed, sleep as a crucial survival-related behavior, is regulated through the hypothalamic circuits, and investigators have specifically linked shortened or disturbed sleep to higher cortisol levels, leading to both increased glucose production, decreased glucose utilization, and eventually insulin resistance [20,24]. Short sleep may also increase levels of the inflammatory marker C-reactive protein, which inhibits physiological function of leptin and contributes to weight gain and impaired glycemic control [3,24]. Although less conclusive, a few studies have also shown excessively long sleep (>8 hours) as a risk factor for diabetes [4,14]. In a two-year prospective study individuals whose sleep duration increases ≥2 hours per night are at an enhanced risk of diabetes [25]. Prolonged sleep elevates inflammatory markers similarly to shortened sleep, resulting in dysfunctional regulation of leptin and food intake [14]. As sleep disturbance is associated with poor glycemic control, optimizing sleep may improve overall diabetes management [26].

On the other hand, some have cautioned that the relationship between sleep and diabetes may be accounted for by other confounding or risk factors. Indeed, both shorter and longer than normal sleep has been documented as core symptoms of depression [27–31]. Higher rates of depression have been consistently found among those diagnosed with diabetes and vice-versa, indicating a bidirectional etiological relationship [28,32,33]. One prospective study of U.S. military members in the Millenium Cohort Study found that, after controlling for baseline covariates, the relationship between depression and diabetes appeared to be insignificant [20]. However, distinct demographics and life experiences of enlisted servicemen might have contributed to these contrasting results. Here, we found that both altered sleep duration and depression were associated with greater odds of a diabetes diagnosis. In particular, the regression models with interaction between sleep and depression did not provide a better fit, suggesting that sleep hours and depression frequency contributed to diabetes risk in a concerted manner. Further, as discussed in detail in the next section, although men relative to women demonstrated a more significant association of both depression and altered sleep hours with the diagnosis of diabetes, both showed a similar pattern of the two risk factors in relation to diabetes. The latter findings suggest an inter-related mechanism by which depression and sleep disturbance contribute to diabetes.

### Sex differences in sleep disturbance and depression as risk factors of diabetes

The effect of sleep on diabetes risk has also been found to vary between sexes. A systematic search of 10 studies found a higher diabetes risk for men sleeping less than 5-6 hours per night, as compared to women [3]. The afore-mentioned study of U.S. military service members reported that, whereas non-depressed women were more likely to be diagnosed with diabetes than non-depressed men, this discrepancy became insignificant when both women and men had depression [20]. Further, Hein et. al, reported that among adults with major depression, only male sex was a significant risk factor for diabetes [28]. Thus, the current finding that aberrant sleep duration – whether too short or long – might impact the risk for diabetes in men more significantly than in women is consistent with this literature. On the other hand, because women as compared to men were more likely to experience and report depression [8], we hypothesized a stronger association between depression and diabetes in women than in men. One possibility for the current finding is a ceiling effect of depression masking the relationship between depression and diabetes in women. However, our data did not support this hypothesis, as women did not appear to show more severe depression in the current data set.

The exact mechanism behind this sex-specific association is not clear. But it should be noted that disturbed sleep has been shown to interfere with testosterone production in men with type 2 diabetes. Given the benefits of testosterone on glucose metabolism, deficient sleep might conceivably lead to poorer glycemic control [34]. Further, given the nature of self-report in the current data, the relationship between sex, depression, and diabetes remains to be investigated [33,35]. Studies with objective measures of glycemic control and sleep quality and duration would be particularly useful in addressing this question.

### Limitations and conclusions

A number of limitations should be considered. First, the data comprised self-reported measures, including sleep duration and subjective experience of depression. Further, as no medical measure of the severity of diabetes was available, the status of diabetes was investigated as a categorical variable. The quality of the data set thus presents a major limitation and the current findings would need to be replicated. Second, these were cross-sectional, observational data; thus, we cannot draw any causal inferences from the current findings. Future studies in a controlled and longitudinal setting ought to examine the direction of the relationship between diabetes, sleep, and depression and how the relationships vary between males and females.

To conclude, the study supports a potential sex-related association between depression frequency and nightly sleep duration and diabetes risk and adds to rapidly growing literature linking mental and physical health. Our findings that both short and long sleep and depression elevate the odds of diabetes support an inter-related rather than independent mechanism of depression and sleep disturbance as risk factors of diabetes. This diabetes risk appears to be stronger in men than in women, suggesting the need of more research to understand sex-specific physiological and psychological processes underlying the care and management of those with or at risk of diabetes.

## Data Availability

This study has used National Health Interview Survey data available at (https://nhis.ipums.org/nhis/).

## Acknowledgements

We thank Dr. Benjamin Baumer and Dr. Bea Capistrant for their advice on the early phase of the study.

## Declarations

### Funding

No funding was received to assist with the preparation of this manuscript.

### Competing interests

The authors have no relevant financial or non-financial interests to disclose. Ethics approval: Not applicable.

### Consent

Not applicable.

## Notes

### Competing Interest Statement

The authors have declared no competing interest.

### Author Declarations

The source data were openly available before the initiation of the study and can be downloaded at https://nhis.ipums.org/nhis/

