## Supplement for "Sex differences in depression and sleep disturbance as inter-related risk factors of diabetes"

### 1 Supplementary Figures and Tables

#### 1.1 Supplementary Figures

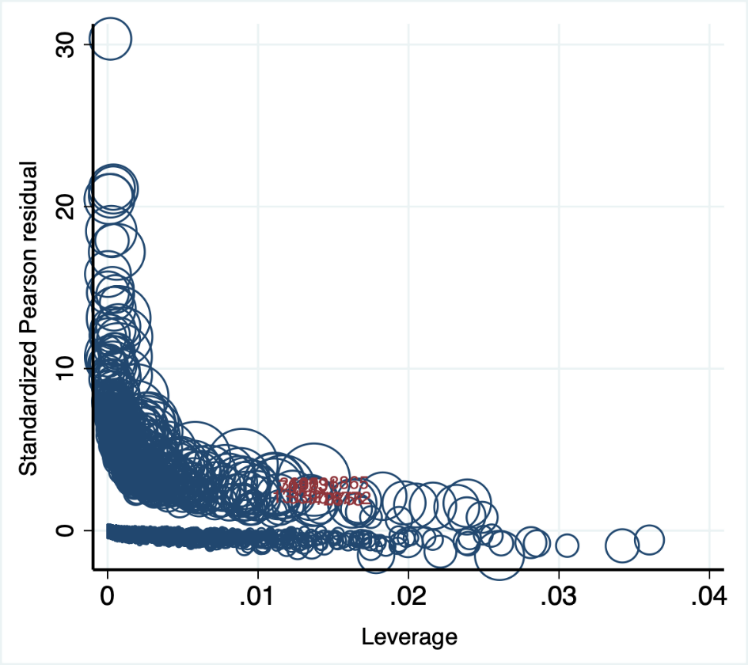

**Supplementary Figure S1.** Standardized Pearson residuals are plotted against Pregibon leverage, with the size of circle proportional to Pregibon delta beta. The data points with standardized Pearson residual >2 AND leverage >0.01 AND Pregibon delta beta >0.01 are marked (subject number: 1009, 2818, 3685, 7023, 8390, 8865, 13152, 17075, 18772, 19731).

**Supplementary Table S1.** Multivariate logistic regression in a sample that excluded 10 subjects as potential outliers (subject number: 1009, 2818, 3685, 7023, 8390, 8865, 13152, 17075, 18772, 19731)

| Variable | Coefficient (SE) | Odds ratio (SE) | p-value |
| --- | --- | --- | --- |
| Sleep | | | |
| ≤5 | 0.44 (0.15) | 1.55 (0.25) | 0.005 |
| 6-7 | 0.36 (0.13) | 1.44 (0.19) | 0.006 |
| 7-8 | Reference | | |
| >8 | 0.44 (0.15) | 1.56 (0.24) | 0.004 |
| Depression | | | |
| Rarely/never | Reference | | |
| Monthly | 0.75 (0.29) | 2.12 (0.67) | 0.011 |
| Often | 0.97 (0.18) | 2.65 (0.51) | <0.001 |
| sex | -0.33 (0.13) | 0.71 (0.08) | 0.008 |
| Depression × sex | | | |
| D(rarely/never)s(W) | Reference |  |  |
| D(monthly)s(M) | -0.33 (0.13) | 0.71 (0.09) | 0.008 |
| D(monthly)s(W) | 0.75 (0.29) | 2.12 (0.62) | 0.011 |
| D(often)s(M) | -0.03 (0.27) | 0.97 (0.27) | 0.907 |
| D(often)s(W) | 0.97 (0.18) | 2.7 (0.49) | <0.001 |
| D(rarely/never)s(W) | 0.32 (0.18) | 1.37 (0.25) | 0.074 |
| Age | 0.05 (0.004) | 1.05 (0.004) | <0.001 |
| BMI | 0.08 (0.006) | 1.07 (0.007) | <0.001 |
| PA | 0.0007 (0.0001) | 1.00 (0.0001) | <0.001 |
| Caucasian | Reference |  |  |
| Asian | 0.74 (0.24) | 2.10 (0.51) | 0.002 |
| African American | 0.58 (0.13) | 1.79 (0.24) | <0.001 |
| Native American | 0.87 (0.32) | 2.39 (0.76) | 0.006 |
| Income |  |  |  |
| low | Reference |  |  |
| Medium | -0.54 (0.13) | 0.58 (0.07) | <0.001 |
| High | -0.95 (0.16) | 0.38 (0.06) | <0.001 |

**Supplementary** **Table S2**. Predictive margins of depression at different levels of sleep hours on the probability of diabetes (estimated using Model 1)

| Depression × sleep | Margin (SE) | p-value |
| --- | --- | --- |
| D(rarely/never)S(≤5 hours) | 0.02 (0.003) | <0.001 |
| D(rarely/never)S(6-7 hours) | 0.02 (0.002) | <0.001 |
| D(rarely/never)S(7-8 hours) | 0.01 (0.001) | <0.001 |
| D(rarely/never)S(>8 hours) | 0.02 (0.002) | <0.001 |
| D(monthly)S(≤5 hours) | 0.03 (0.006) | <0.001 |
| D(monthly)S(6-7 hours) | 0.02 (0.004) | <0.001 |
| D(monthly)S(7-8 hours) | 0.04 (0.007) | <0.001 |
| D(monthly)S(>8 hours) | 0.04 (0.006) | <0.001 |
| D(often)S(≤5 hours) | 0.04 (0.005) | <0.001 |
| D(often)S(6-7 hours) | 0.04 (0.005) | <0.001 |
| D(often)S(7-8 hours) | 0.03 (0.004) | <0.001 |
| D(often)S(>8 hours) | 0.04 (0.005) | <0.001 |
